# Does COVID-19 vaccination improve mental health? A difference-in-difference analysis of the Understanding Coronavirus in America study

**DOI:** 10.1101/2021.07.19.21260782

**Authors:** Jonathan Koltai, Julia Raifman, Jacob Bor, Martin McKee, David Stuckler

**Affiliations:** Assistant Professor, Department of Sociology, University of New Hampshire, Durham, US; Assistant Professor, Department of Health Law, Policy, and Management, Boston University, School of Public Health, Boston, Massachusetts; Assistant Professor, Department of Global Health, Boston University School of Public Health, Boston, Massachusetts; Full Professor, Centre for Global Chronic Conditions, London School of Hygiene and Tropical Medicine, London, UK; Full Professor, Dondena Centre for Research on Social Dynamics and Public Policy, Bocconi University, Milan, Italy

## Abstract

**Background:** Mental health problems increased during the COVID-19 pandemic. Knowledge that one is less at risk after being vaccinated may alleviate distress, but this hypothesis remains unexplored. Here we test whether psychological distress declined in those vaccinated against COVID-19 in the US and whether changes in perceived risk mediated any association.

**Methods:** A nationally-representative cohort of U.S. adults (N=5,792) in the Understanding America Study were interviewed every two weeks from March 2020 to June 2021 (28 waves). Difference-in-difference regression tested whether getting vaccinated reduced distress (PHQ-4 scores), with mediation analysis used to identify potential mechanisms, including perceived risks of infection, hospitalization, and death.

**Results:** Vaccination was associated with a 0.09 decline in distress scores (95% CI:-0.15 to -0.04) (0-12 scale), a 5.7% relative decrease compared to mean scores in the wave prior to vaccination. Vaccination was also associated with an 8.44 percentage point reduction in perceived risk of infection (95% CI:-9.15% to -7.73%), a 7.44-point reduction in perceived risk of hospitalization (95% CI:-8.07% to -6.82%), and a 5.03-point reduction in perceived risk of death (95% CI:-5.57% to -4.49%). Adjusting for risk perceptions decreased the vaccination-distress association by two-thirds. Event study models suggest vaccinated and never vaccinated respondents followed similar PHQ-4 trends pre-vaccination, diverging significantly post-vaccination. Analyses were robust to individual and wave fixed effects, time-varying controls, and several alternative modelling strategies. Results were similar across sociodemographic groups.

**Conclusion:** Receiving a COVID-19 vaccination was associated with declines in distress and perceived risks of infection, hospitalization, and death. Vaccination campaigns could promote these additional benefits of being vaccinated.

## Introduction

On May 13^th^ 2020, the United Nations warned that although COVID was primarily an infectious disease, it was also sowing the “seeds of a major mental health crisis”(1). Surveys in the United States reveal elevated levels of psychological distress, anxiety, and suicidal ideation since the onset of the pandemic.(2–6) Several factors have contributed to these findings, including loss of income and work, food insecurity, social isolation, additional care-giving burdens, substance use, and racialized discrimination.(7–18) Personal experience of COVID-related illness and death or hospitalization of a loved one may also contribute.(19, 20) One recent study points to anticipatory fears, with perceived risk of infection and mortality explaining 20.7% of the increased distress between March and June 2020.(21)

High rates of vaccination are crucial to prevent the spread of COVID-19 and its many consequences, including worsening mental health. Yet, as of July 4^th^, only 67.1% percent of US adults have been vaccinated,(22) falling short of President Biden’s target of 70% by the same date, and daily vaccination rates have fallen sharply.(23) While side effects and safety top the list of concerns of those not vaccinated, lack of information and access remain barriers for vulnerable individuals, particularly people of color.(24) Many people of color who are not yet vaccinated express vaccine hesitancy but also high perceived risk from COVID-19 infection.(25) Meanwhile, although the poor and those facing food and housing insecurity are less likely to be vaccinated, many, especially those with children, want a vaccine.(26)

Are there individual and social benefits of vaccination beyond preventing infection? One hypothesis, so far unexplored, is whether vaccination improves mental health by reducing anticipatory fears of infection, hospitalization, and death. We use a difference-in-difference method with nationally-representative longitudinal data to test whether vaccination for COVID-19 reduces psychological distress and, if so, whether lower perceived risk mediates this association.

## Study Design and Methods

### Data Source

We used data from the Understanding Coronavirus in America (UCA) study,(27) an extension of an internet-based, nationally representative longitudinal survey.

We examined 28 survey waves, between March 2020 and June, 2021. We restricted the sample to participants in at least 2 survey waves with non-missing values for our exposure, outcome, and covariates and excluded person-period observations in which respondents indicated being unsure of their vaccination status. Our primary analytic sample included 5,792 individuals. Table 1A in the supplement shows dates of each study wave.

### Exposure

Our primary exposure was vaccination status, coded as 1 beginning in the first wave in which the respondent answered yes to the question “Have you gotten vaccinated for the coronavirus?” and imputed as 1 thereafter. It was coded as 0 for “No” and imputed as 0 in each period prior to Wave 21 (December 23, 2020 to January 18, 2021), the first time this question was asked.

### Outcome

Psychological distress was assessed using the Patient Health Questionnaire (PHQ-4) developed by Kroenke et al.(28), and validated by Lowe et al.(29) The scale consists of the first two items from the PHQ-9 and the Generalized Anxiety Disorder-7 (GAD-7) which assess core criteria for depressive (e.g. “Little interest or pleasure in doing things”) and anxiety disorders (e.g. “Feeling nervous, anxious or on edge”) respectively. Participants report how often they have been bothered by these problems over the last 2 weeks on a four-point scale scored as 0 (“not at all”), 1 (“several days”), 2 (“more than half the days”), or 3 (“nearly every day”). Scores on the scale range from 0 to 12 with higher scores indicating greater distress. Our main analyses use total PHQ-4 scores. We also use a variable for severe distress in supplementary analyses, coded 1 if PHQ-4 scores 9 or above and 0 for those below.(28)

### Mediators

Participants were asked “On a scale from 0 to 100%, what is the chance that you will get the coronavirus in the next three months?” and then “If you do get the coronavirus, what is the percent chance you will be hospitalized (spend at least one night in the hospital) from it?” Finally, perceived infection-fatality risk was assessed by asking “If you do get infected with the coronavirus, what is the chance you will die from it?”, with responses also recorded as 0 to 100%.

### Covariates

In our main analyses we included individual fixed effects (separate indicators for each person) to adjust for all time-invariant respondent characteristics. We included survey wave fixed effects (separate indicators for each wave) to adjust for national secular trends in mental health. We also adjusted for several time-varying, self-reported covariates: receiving Supplemental Nutrition Assistance Program (SNAP) benefits in the month prior to the survey, receiving unemployment insurance past 14 days, and employment status at the time of the survey.

### Statistical Analysis

We first described the demographic characteristics of the sample by vaccination status, which are shown in Table 2A of the supplement. We then use graphical methods to visualize unadjusted temporal trends in psychological distress scores (Figure 1A) and perceived risk factors (Figure 2A) in the supplementary appendix.

Next, we use two-way fixed effects models to assess the association between receiving a COVID-19 vaccination and changes in mental health. These models take the form:

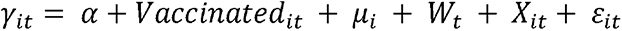

Where *γ_it_* denotes psychological distress for individual i at wave t. *μ_i_* and *W_t_* are individual and wave fixed effects respectively, *X_it_* represents time-varying controls. *Vaccinated_it_* is an indicator variable that switches to 1 in first wave in which the respondent answered yes to the question “Have you gotten vaccinated for the coronavirus?” and remains as 1 thereafter.

To assess how the outcomes of interested changed over time before and after vaccination, we estimated so-called “event study” models, wherein *Vaccinated_it_* was replaced with dummy variables indicating the number of periods prior to or following a respondent’s first report of COVID-19 vaccination.(30, 31) We binned dummy variables for lags at 4 or more waves (8 or more weeks) post-vaccination, reflecting the 99^th^ percentile of the distribution of observations in our sample. The event study specification provides two important pieces of information not observable in the single-coefficient two-way fixed effects model.(32) First, that model assumes that distress would have continued along the same trajectory in those who were or were not vaccinated. While this cannot be tested explicitly, including all event leads reveals in the pre-treatment period when coefficients for leads (pre-vaccination) differing significantly from zero would suggest violation of the parallel trends assumption. By contrast, non-zero coefficients for lags (post-vaccination) indicate statistically significant treatment effects. Second, the lags make it possible to see whether the effects grow or shrink over time, and whether they persist. For all analyses, we clustered standard errors by individuals to account for serial correlation.

We conducted several sensitivity checks to test whether the effects of vaccination vary by demographic subgroups, and whether they are robust to alternative modelling strategies.

## Results

Table 2A in the supplement shows a gradient in vaccination status by age, income, and education, with higher rates of vaccination observed among older respondents and those at the higher end of the socioeconomic status spectrum. Among self-reported race categories, Asians had the highest rates of vaccination (69%), followed by Hawaiian/Pacific Islander (59%), Whites (57%), mixed race respondents (49%), Black respondents (48%), and American Indian and Alaska Natives (38%). Males had slightly higher rates of vaccination (59%) compared to females (54%) in our sample. Married individuals living with partners had the highest rates of vaccination (61%) among marital status groups, while separated individuals had the lowest (36%).

Figure 1A shows the secular trends in distress scores for never vaccinated respondents and respondents who became vaccinated during our study period. Distress scores increase for both groups at the outset of the pandemic, peaking in Wave 2 (4/1/2020 to 4/27/2020), then decline steadily until Wave 7 (6/10/2020 to 7/6/2020). Notwithstanding some fluctuations, distress scores then remain relatively stable for both groups until Wave 26 (3/17/2021 to 4/27/2021), after which distress declines slightly for respondents who became vaccinated and increases slightly for never vaccinated respondents. Wave 26 corresponds to the median wave of vaccination in our sample.

Figure 2A shows secular trends in risk perceptions for never vaccinated respondents and respondents who became vaccinated during our study period. Here, while respondents who became vaccinated exhibited slightly higher levels of risk perceptions for the majority of the study period, both groups shared similar trends until Wave 25 (2/17/2021 to 3/29/2021), corresponding to the wave prior to the median wave of vaccination in our sample. Risk perceptions then decline for respondents who became vaccinated, falling below the mean risk perception levels of respondents who did not become vaccinated.

Table 1 shows our difference-in-difference estimates for the association between vaccination and perceived risk of infection in Model 1, perceived risk of hospitalization in Model 2, and perceived risk of death in Model 3. All models adjust for individual and wave fixed effects, and time-varying unemployment insurance, SNAP, and employment status. Here, vaccination is associated with an 8.44 percentage point reduction in perceived risk of infection (95% CI: -9.15% to -7.73%), a 7.44-point reduction in perceived risk of hospitalization (95% CI: -8.07% to -6.82%), and a 5.03-point reduction in perceived risk of death (95% CI: -5.57% to - 4.49%).

**Table 1.**
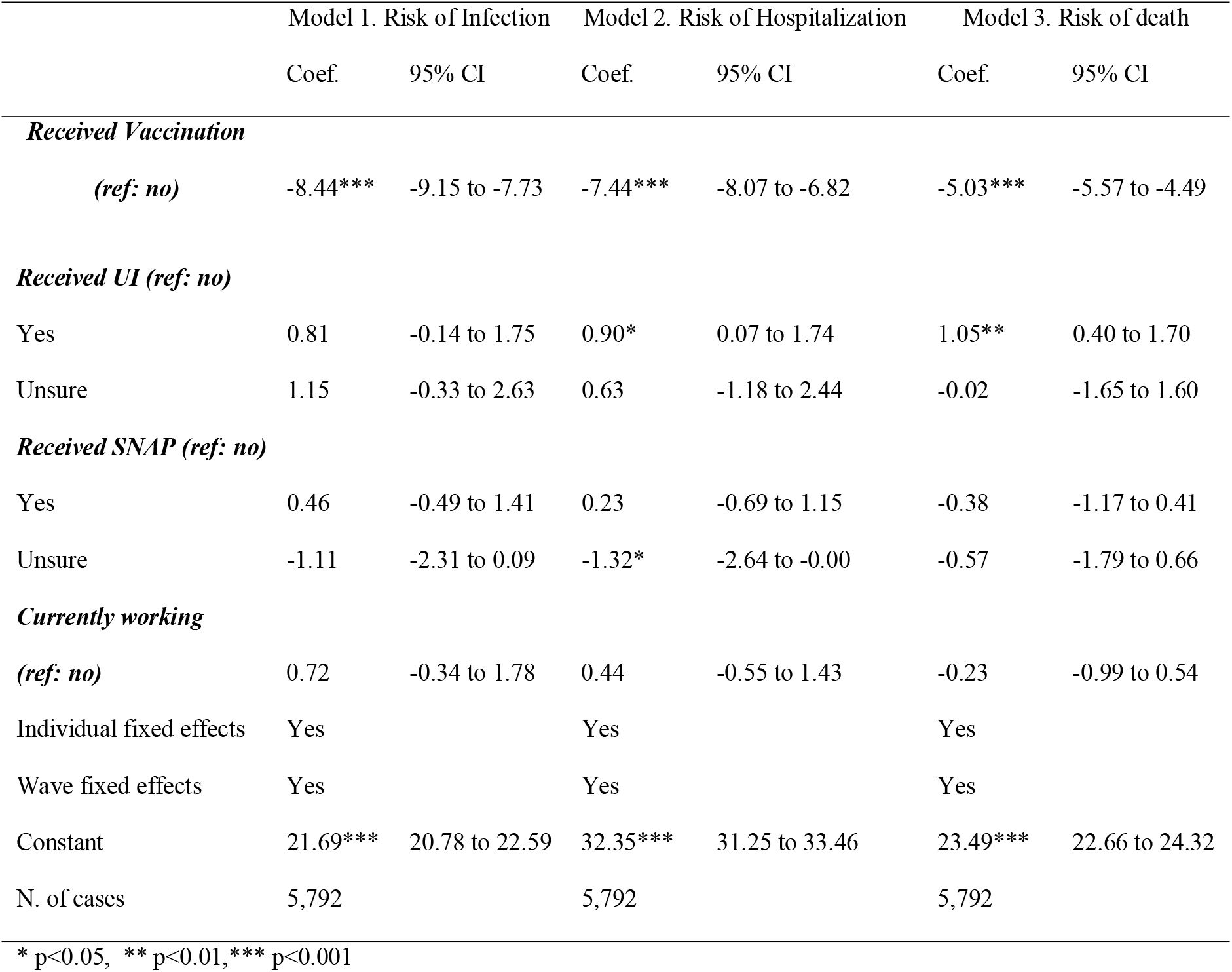
Two-way fixed effects models with perceived risk factors regressed on vaccination status, April 2020 to June 2021 (N= 5,792)

Table 2 shows our primary difference-in-difference analyses. In Model 1, receiving vaccination is associated with a -.09 decrease in distress scores (p < 0.001; 95% CI: -0.15 to - 0.04), and this relationship remains unchanged after adjusting for unemployment insurance, SNAP, and employment status in Model 2. This estimate corresponds to a 5.7% relative reduction in distress compared to mean distress scores in the wave prior to vaccination.

**Table 2.**
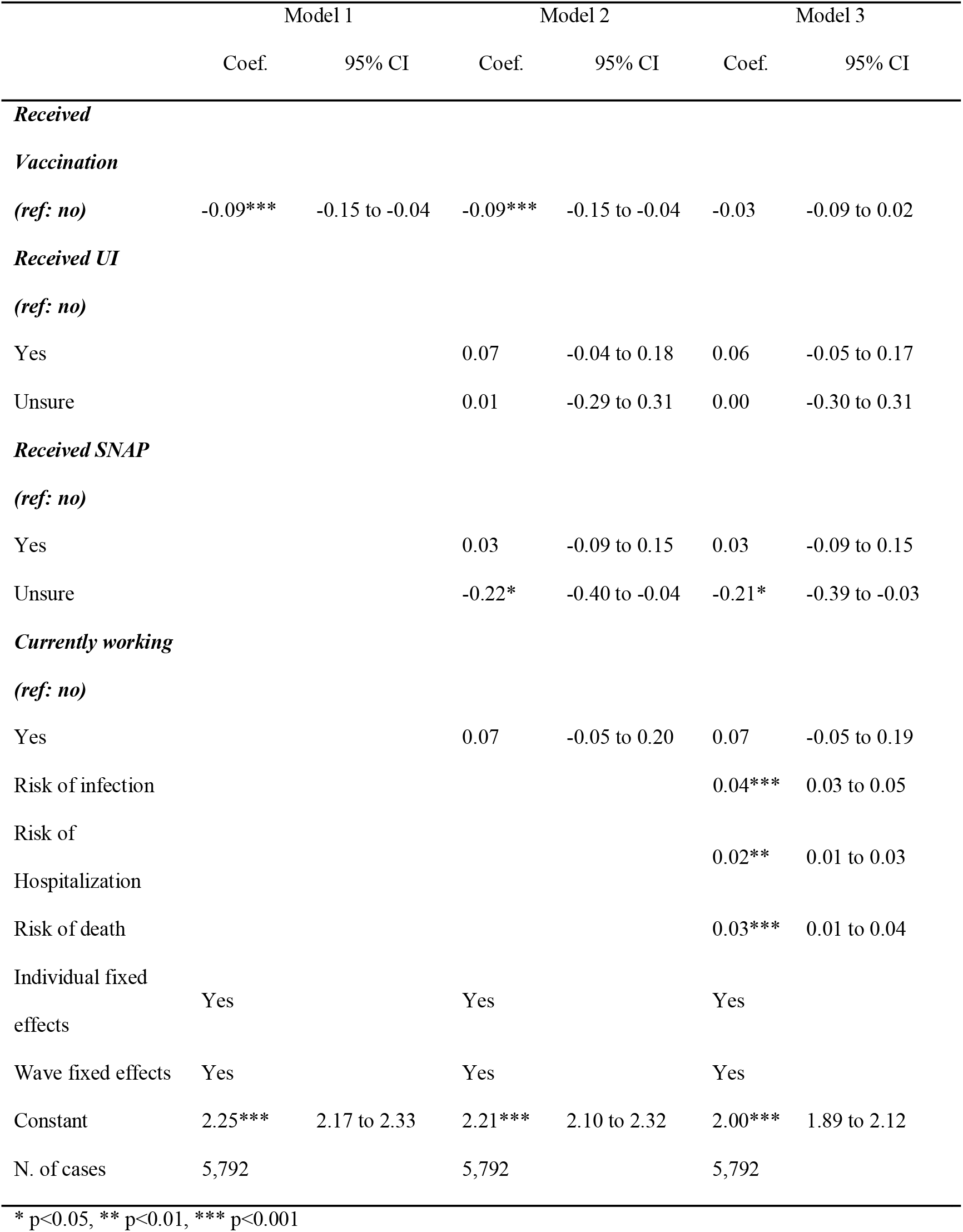
Two-way fixed effects models with psychological distress (PHQ-4) regressed on vaccination status and perceived risk factors, April 2020 to June 2021 (N= 5,792) **Notes:** Standard errors are clustered at the individual level. Coefficients for perceived risk factors are expressed as a 10-percentage point increase.

Model 3 adjusts for risk perceptions, which are all independently associated with distress net of each other. A 10-percentage point increase in perceived risk of infection is associated with a 0.04 increase in psychological distress scores (p <0.001; 95% CI: 0.03 to 0.05); perceived risk of hospitalization is associated with a 0.02 increase in psychological distress scores (p <0.01; 95% CI: 0.01 to 0.03); death is associated with a 0.03 increase in psychological distress scores (p <0.001; 95% CI: 0.01 to 0.04). Importantly, adjusting for risk perceptions in Model 3 reduces the coefficient for vaccination by two-thirds, to -0.03, which is no longer statistically significant (p = 0.22; 95% CI: -0.09 to 0.02). Taken together, these models suggest that the receiving the COVID-19 vaccination reduces psychological distress, and that this effect is transmitted largely through declines in perceived risk of infection, hospitalization, and death.

The event study analyses in Figures 1 and 2 provide additional support for our findings in Table 2. Figure 1 shows vaccinated and never vaccinated respondents followed similar trends in distress scores prior to vaccination, and that these diverged significantly afterwards, with vaccinated respondents experiencing significant declines in distress. Importantly, these treatment effects persist for 8 weeks post vaccination. It takes a few waves for the effect to be fully realized, which suggests that our regression estimates likely underestimate the true effect. In Figure 2 we observe slight differences in risk perceptions between vaccinated and never vaccinated respondents in the pre-treatment period, although these are stable over time. Following vaccination, these trends diverge, with vaccinated individuals experiencing large reductions risk perceptions relative to those who were never vaccinated in our sample. Finally, Figure 1A (supplement) shows the event study results for the effect of vaccination on distress after adjusting for risk perceptions. Consistent with Model 3 in Table 2, we observe no significant treatment effects of vaccination after accounting for changing risk perceptions associated with vaccination.

**Figure 1.**
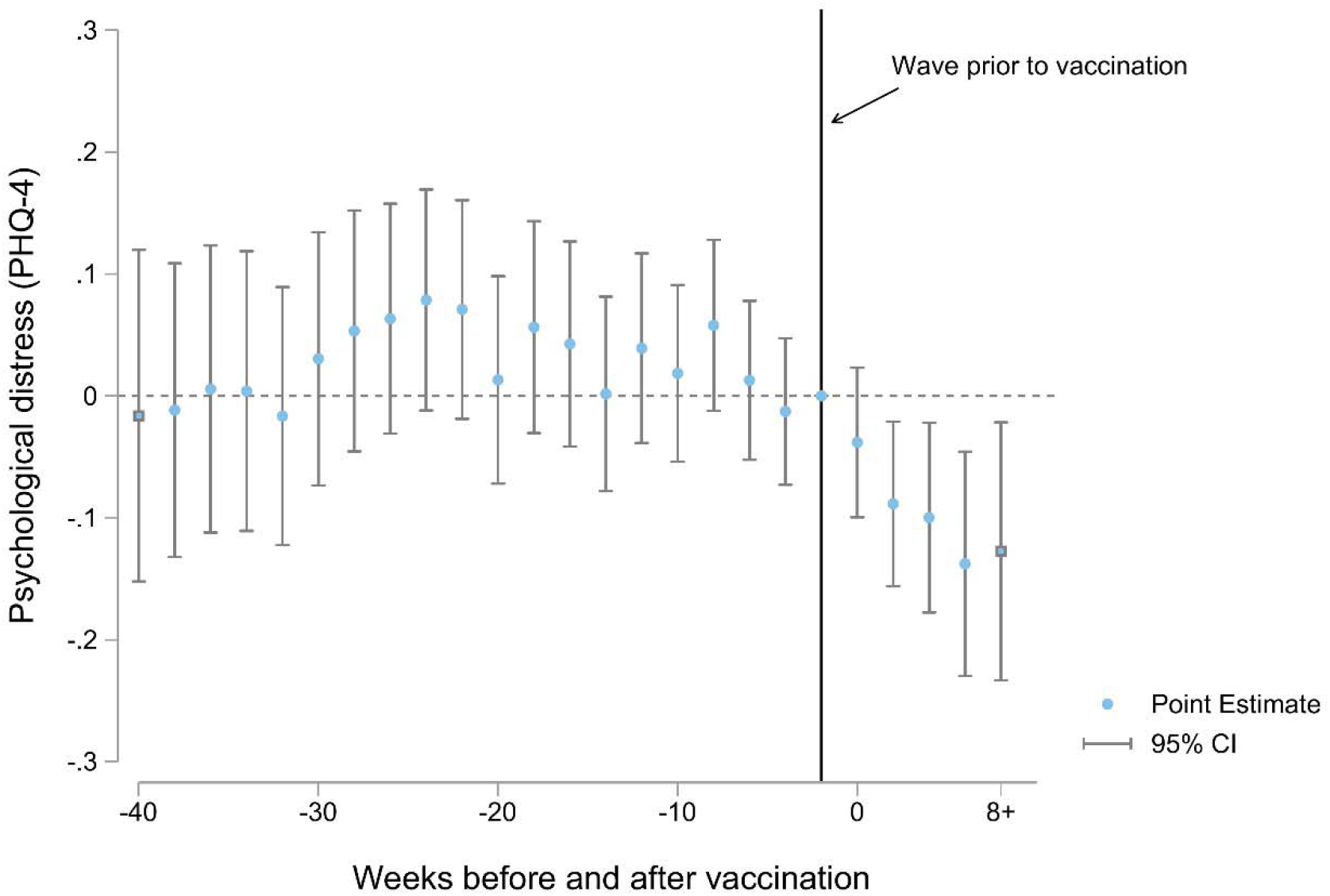
Difference-in-Differences Estimates of the Association Between Receiving the COVID-19 Vaccine and Psychological Distress Notes: Each point estimate refers to the change in distress between vaccinated and never vaccinated individuals, compared to their baseline differential in the wave immediately prior to vaccination. Models control for individual and wave fixed effects, receiving Supplemental Nutrition Assistance Program (SNAP) benefits in the month prior to the survey, whether the respondent received unemployment insurance in the past 14 days, and employment status at the time of the survey.

**Figure 2:**
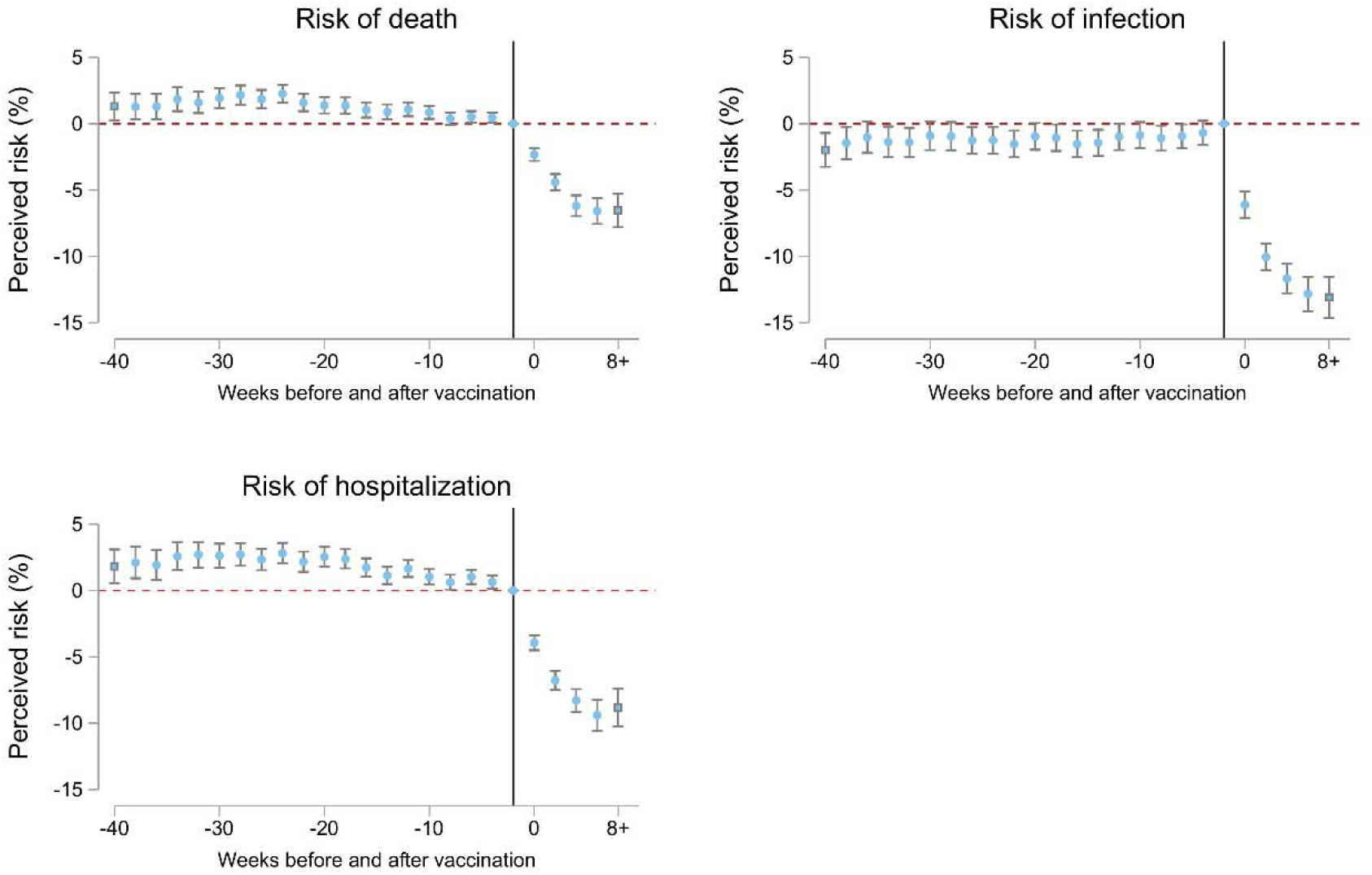
Difference-in-Differences Estimates of the Association Between Receiving the COVID-19 Vaccine and Perceived Risk Factor Notes: Each point estimate refers to the change in risk perceptions between vaccinated and never vaccinated individuals, compared to their baseline differential in the wave immediately prior to vaccination (reference line on the x-axis). Models control for individual and wave fixed effects, receiving Supplemental Nutrition Assistance Program (SNAP) benefits in the month prior to the survey, whether the respondent received unemployment insurance in the past 14 days, and employment status at the time of the survey.

### Sensitivity checks

Figures 4A-8A in the supplement test whether the effects of vaccination vary by demographic subgroups. Each figure represents two-way fixed effects models with psychological distress scores regressed on vaccination status stratified by age groups (Figure 4A), race/ethnicity (Figure 5A), education (Figure 6A), gender (Figure 7A), and household income (Figure 8A). Here, we do not observe substantive differences in the effect of vaccination across sociodemographic groups.

Tables 3A-6A show how the patterns described in our main analyses remain robust after using sample weights, restricting our models to respondents aged 65+, including state-by-wave fixed effects, and when using an indicator for severe psychological (PHQ-4 scores ≥ 9) as the dependent variable. With respect to the latter, we found that vaccination was associated with a 0.6 percentage point reduction in severe distress from a baseline prevalence of 0.04% in the wave prior to vaccination, corresponding to a 15% relative decline.

## Discussion

This study has several important findings. First, vaccination was associated with a 5.7% reduction in PHQ-4 distress scores and a 15% reduction in severe distress (PHQ-4 scores ≥ 9). Vaccination was also associated with an 8.44 percentage point reduction in perceived risk of infection, a 7.44-point reduction in perceived risk of hospitalization, and a 5.03-point reduction in perceived risk of death. Adjusting for risk perceptions decreased the association between vaccination and distress by two-thirds, to statistical insignificance. Complementary panel event study models suggest that vaccinated and never vaccinated respondents shared similar trends in distress prior to vaccination, and that these trends diverged significantly following vaccination.

As with all observational analyses, our study has clear limitations. First, measurements of both the distress and vaccination rely on self-report, which may be biased. Second, the web-based sample may not be truly representative of the US population and individuals from underrepresented racial and ethnic groups. Third, our analysis was designed to capture only the direct effect of an individual becoming vaccinated on their own mental health. However, vaccination is likely to have myriad positive spillover effects on mental health that are not captured in this study. Mental health may improve as friends and family become vaccinated, as the economy rebounds, as community prevalence of virus falls, and as fewer people suffer major illness or death. The respondents in the UCA study likely benefited from vaccine scale-up beyond their own vaccination status. Additionally, their getting vaccinated likely benefited other peoples’ mental health. As a result, our findings likely substantially underestimate the beneficial effect of vaccination for mental health at the population level.

Notwithstanding these limitations, our study has several key strengths. First, to our knowledge, this is the first study to assess the psychological impacts of COVID-19 vaccination. Second, our main findings are robust to time-varying controls, individual and wave fixed effects, and several alternative modelling strategies. Importantly, our analyses were robust to state-by-wave fixed effects, ruling out confounding due to time-varying factors at the state-level, such as rates of infection or policy implementation. Third, our event study models show that the mental health benefits of vaccination persist for at least 8 weeks, suggesting that these effects are not a signal of fleeting relief.

Our results have important public health and policy implications. Murphy and colleagues(35) suggest that messages tailored to vaccine hesitant or resistant individuals could emphasize the personal benefits of vaccination against COVID-19. We provide an evidence base for these personal benefits that underscore mental health improvements following vaccination in a nationally representative sample of U.S. adults. We also demonstrate that vaccination is a key lever in reducing the mental health burdens associated with the COVID-19 pandemic.

## Data Availability

Data are publicly available from the Understanding Coronavirus in America Study.

https://uasdata.usc.edu/index.php

## Acknowledgements

The project described in this paper relies on data from survey(s) administered by the Understanding America Study, which is maintained by the Center for Economic and Social Research (CESR) at the University of Southern California. The content of this paper is solely the responsibility of the authors and does not necessarily represent the official views of USC or UAS. The collection of the UAS COVID-19 tracking data is supported in part by the Bill & Melinda Gates Foundation and by grant U01AG054580 from the National Institute on Aging, and many others. The authors would also like to thank Atheendar Venkataramani for his helpful comments on an earlier version of this manuscript.

## Supplementary Figures and Tables

**Figure 1A:**
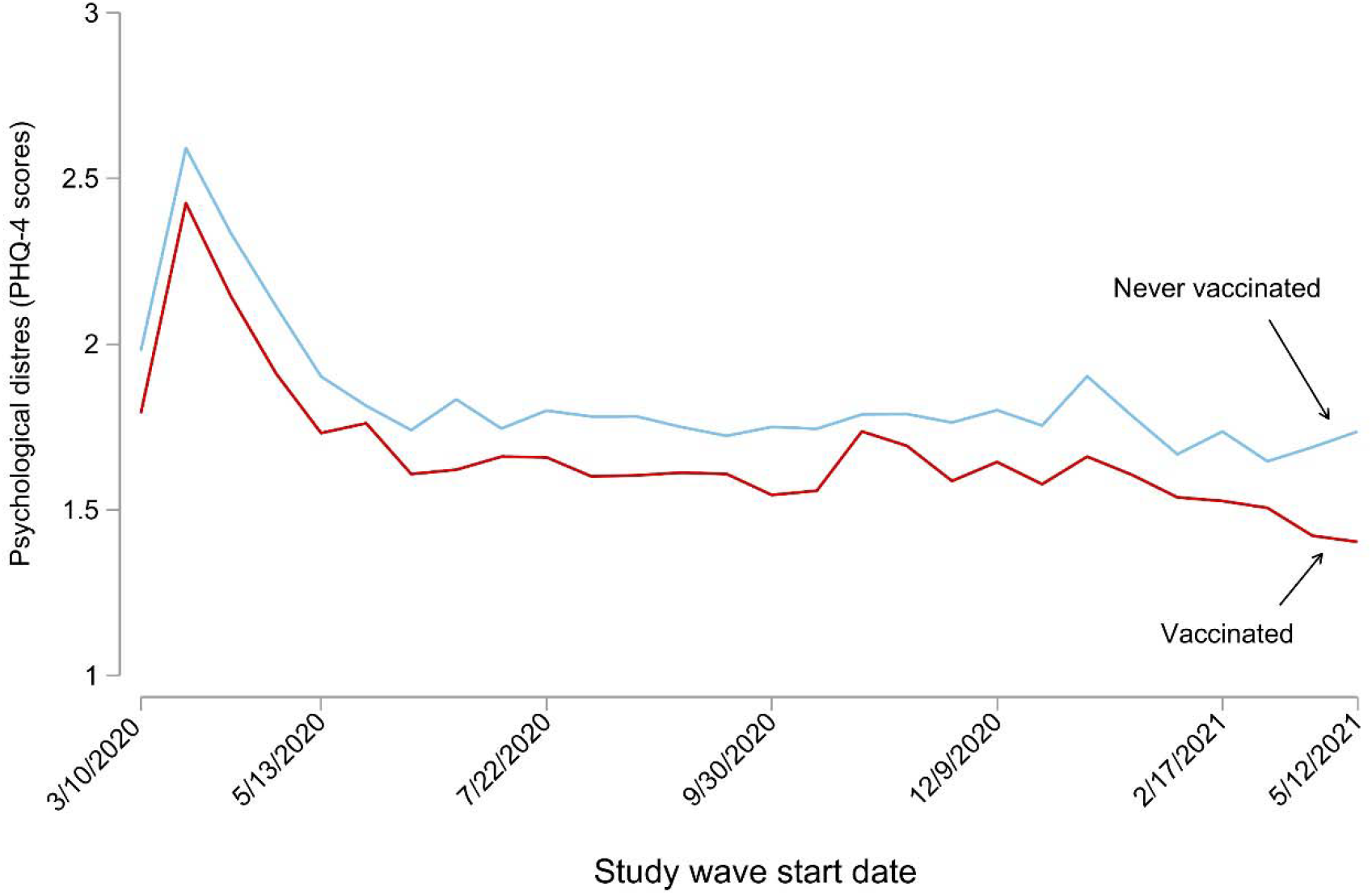
Unadjusted temporal trends in psychological distress over 28 waves in the Understanding Coronavirus in America study, March 2020 to June 2021

**Figure 2A:**
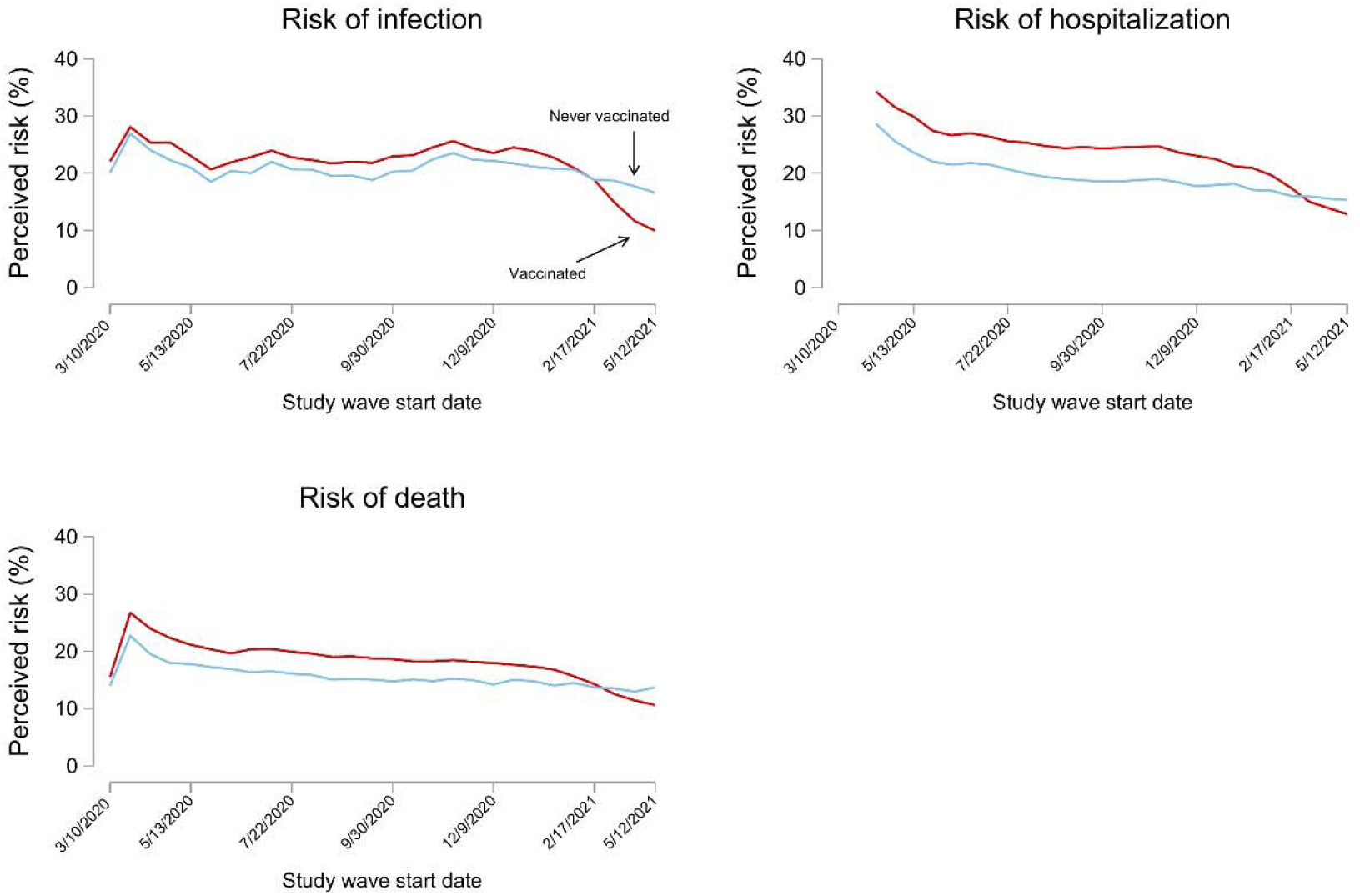
Unadjusted temporal trends in risk perceptions over 28 waves in the Understanding Coronavirus in America study, March 2020 to June 2021 Notes: Red lines are secular trends in risk perceptions over time for respondents who became vaccinated. Blue lines are secular trends over time for respondents who did not become vaccinated.

**Figure 3A:**
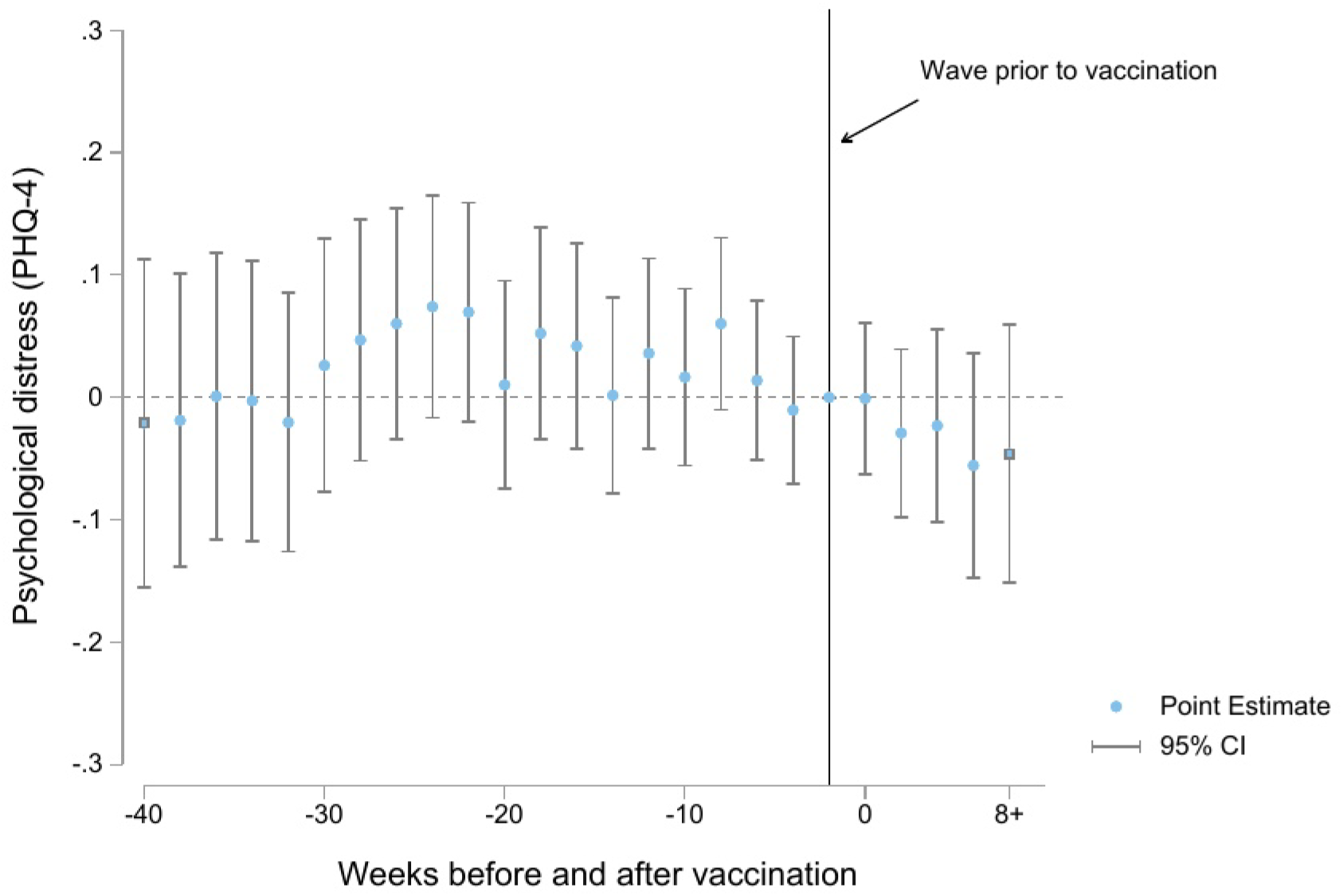
Difference-in-Differences Estimates of the Association Between Receiving the COVID-19 Vaccine and Psychological Distress (**including risk factors as mediators**) Notes: Each point estimate refers to the change in distress between vaccinated and never vaccinated individuals, compared to their baseline differential in the wave immediately prior to vaccination. Models control for individual and wave fixed effects, receiving Supplemental Nutrition Assistance Program (SNAP) benefits in the month prior to the survey, whether the respondent received unemployment insurance in the past 14 days, and employment status at the time of the survey.

**Figure 4A.**
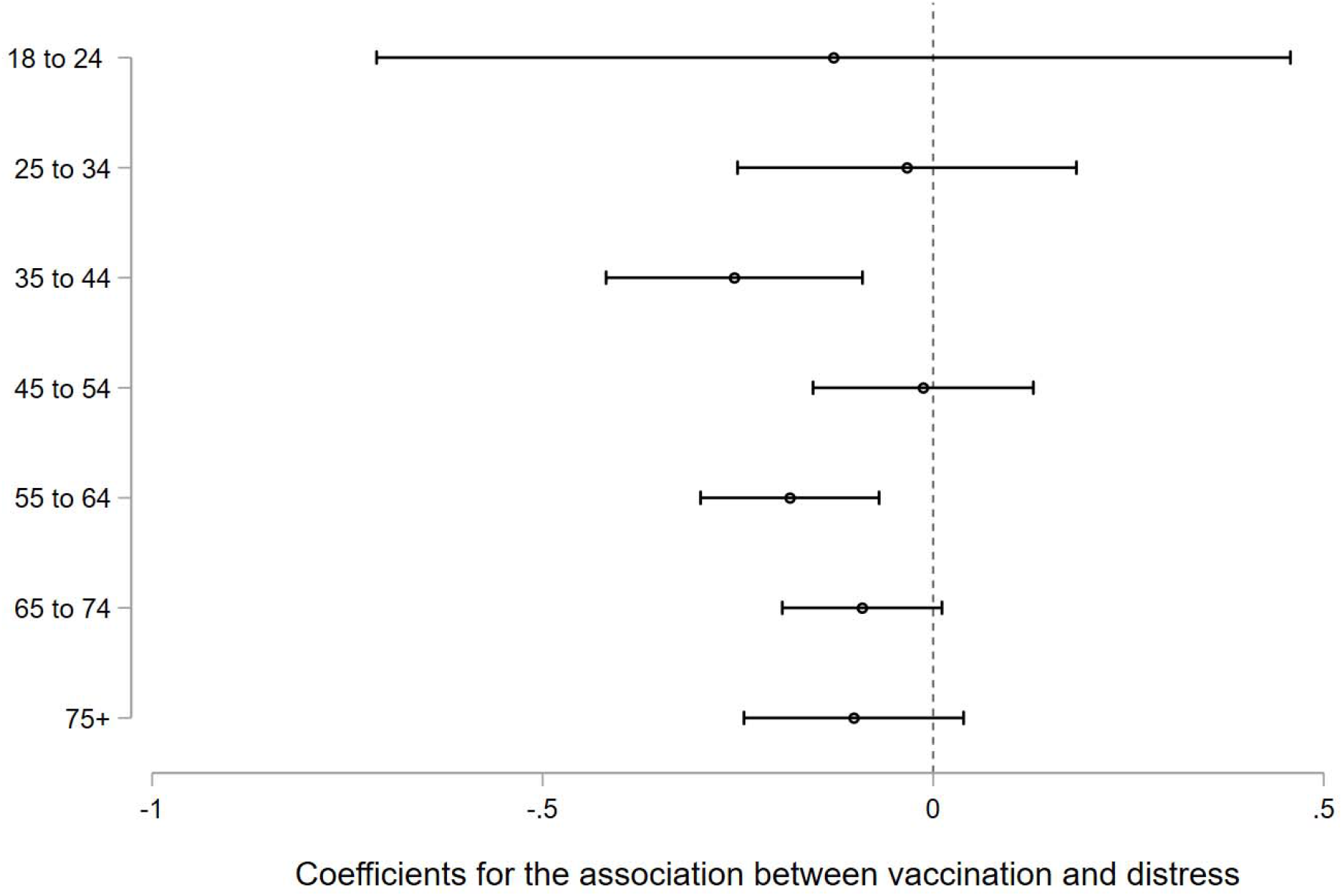
Two-way fixed effects models with psychological distress (PHQ-4) regressed on vaccination status, stratified by age, April 2020 to June 2021 Notes: Point estimates and 95% confidence intervals are from separate two-way fixed effects models stratified by subgroup. Models control for individual and wave fixed effects, receiving Supplemental Nutrition Assistance Program (SNAP) benefits in the month prior to the survey, whether the respondent received unemployment insurance in the past 14 days, and employment status at the time of the survey. Standard errors are clustered at the individual level.

**Figure 5A.**
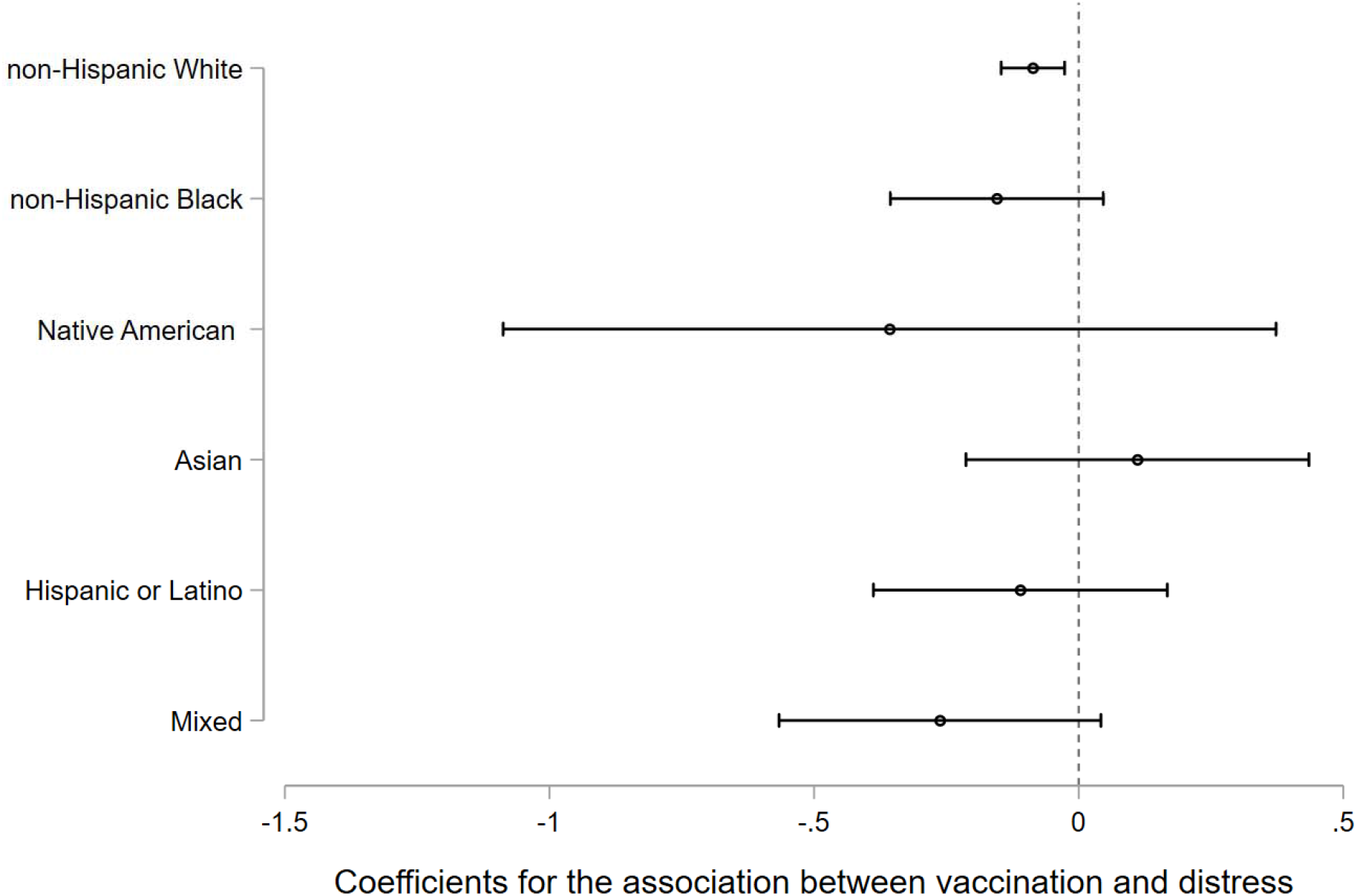
Two-way fixed effects models with psychological distress (PHQ-4) regressed on vaccination status, stratified by self-reported race/ethnicity, April 2020 to June 2021 Notes: Point estimates and 95% confidence intervals are from separate two-way fixed effects models stratified by subgroup. Native Hawaiian or Other Pacific Islander respondents were combined with Asian respondents due to extremely small cell sizes for the former groups. Models control for individual and wave fixed effects, receiving Supplemental Nutrition Assistance Program (SNAP) benefits in the month prior to the survey, whether the respondent received unemployment insurance in the past 14 days, and employment status at the time of the survey. Standard errors are clustered at the individual level.

**Figure 6A.**
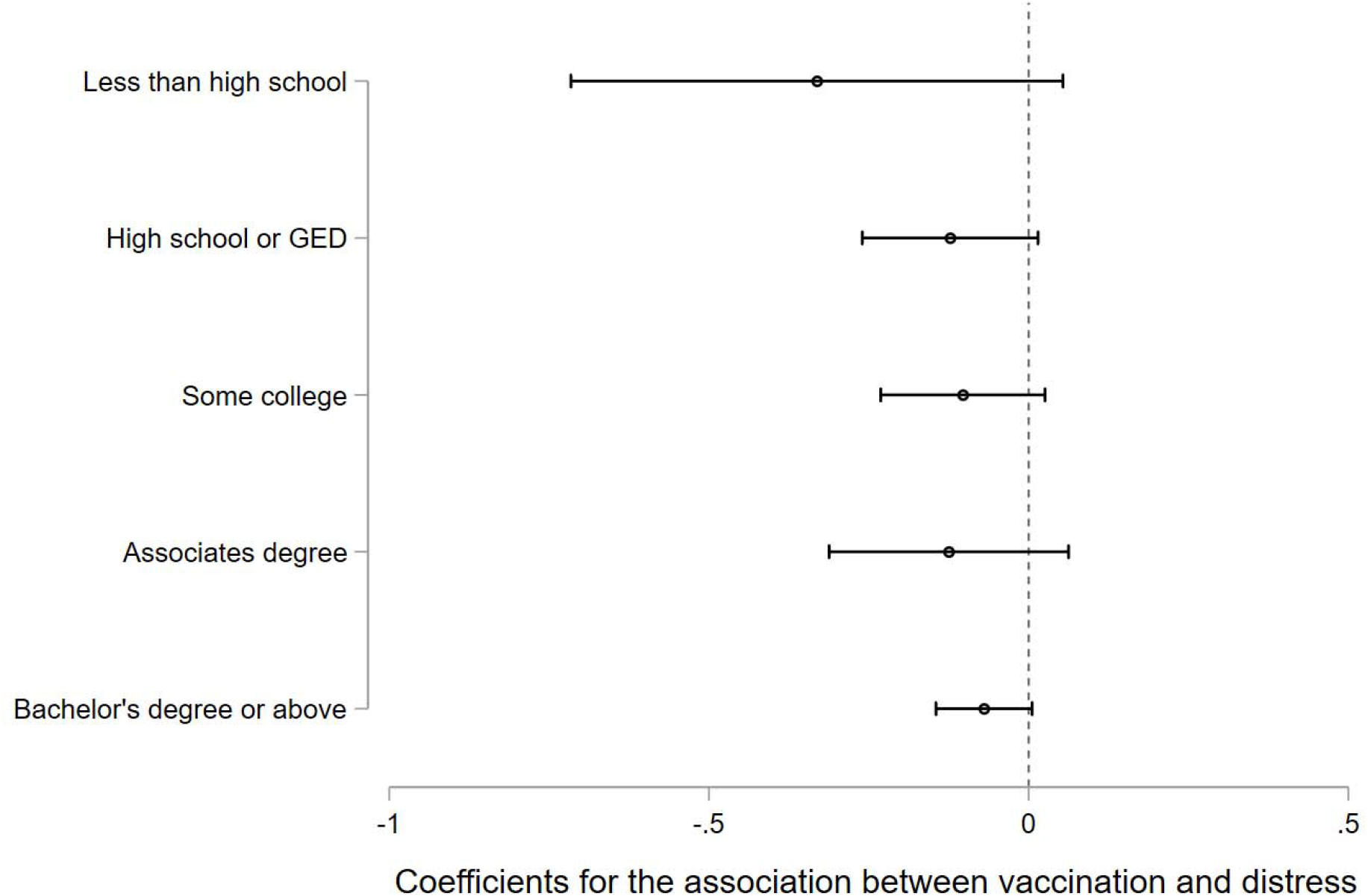
Two-way fixed effects models with psychological distress (PHQ-4) regressed on vaccination status, stratified by education, April 2020 to June 2021 Notes: Point estimates and 95% confidence intervals are from separate two-way fixed effects models stratified by subgroup. Models control for individual and wave fixed effects, receiving Supplemental Nutrition Assistance Program (SNAP) benefits in the month prior to the survey, whether the respondent received unemployment insurance in the past 14 days, and employment status at the time of the survey. Standard errors are clustered at the individual level.

**Figure 7A.**
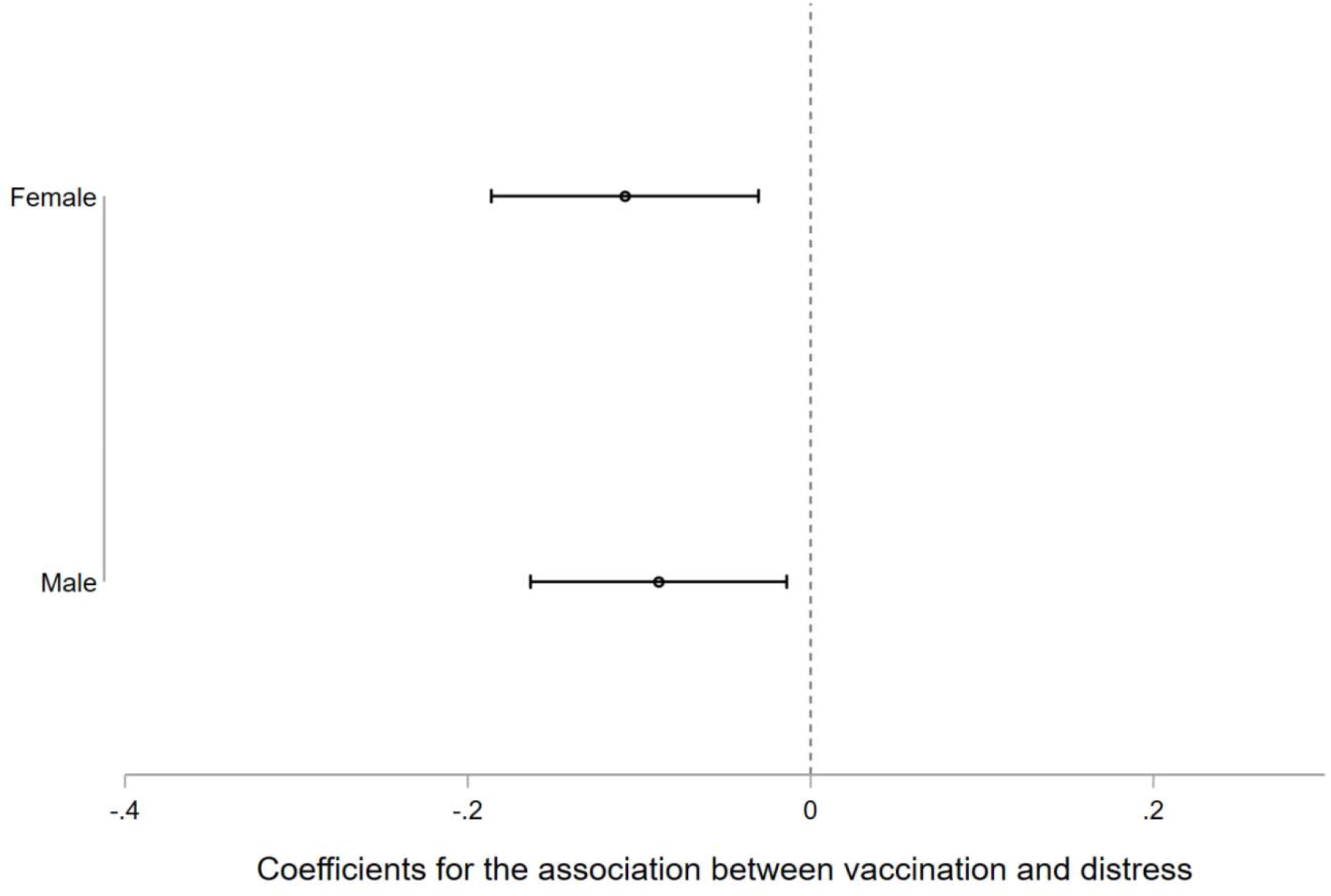
Two-way fixed effects models with psychological distress (PHQ-4) regressed on vaccination status, stratified by gender, April 2020 to June 2021 Notes: Point estimates and 95% confidence intervals are from separate two-way fixed effects models stratified by subgroup. Models control for individual and wave fixed effects, receiving Supplemental Nutrition Assistance Program (SNAP) benefits in the month prior to the survey, whether the respondent received unemployment insurance in the past 14 days, and employment status at the time of the survey. Standard errors are clustered at the individual level.

**Figure 8A.**
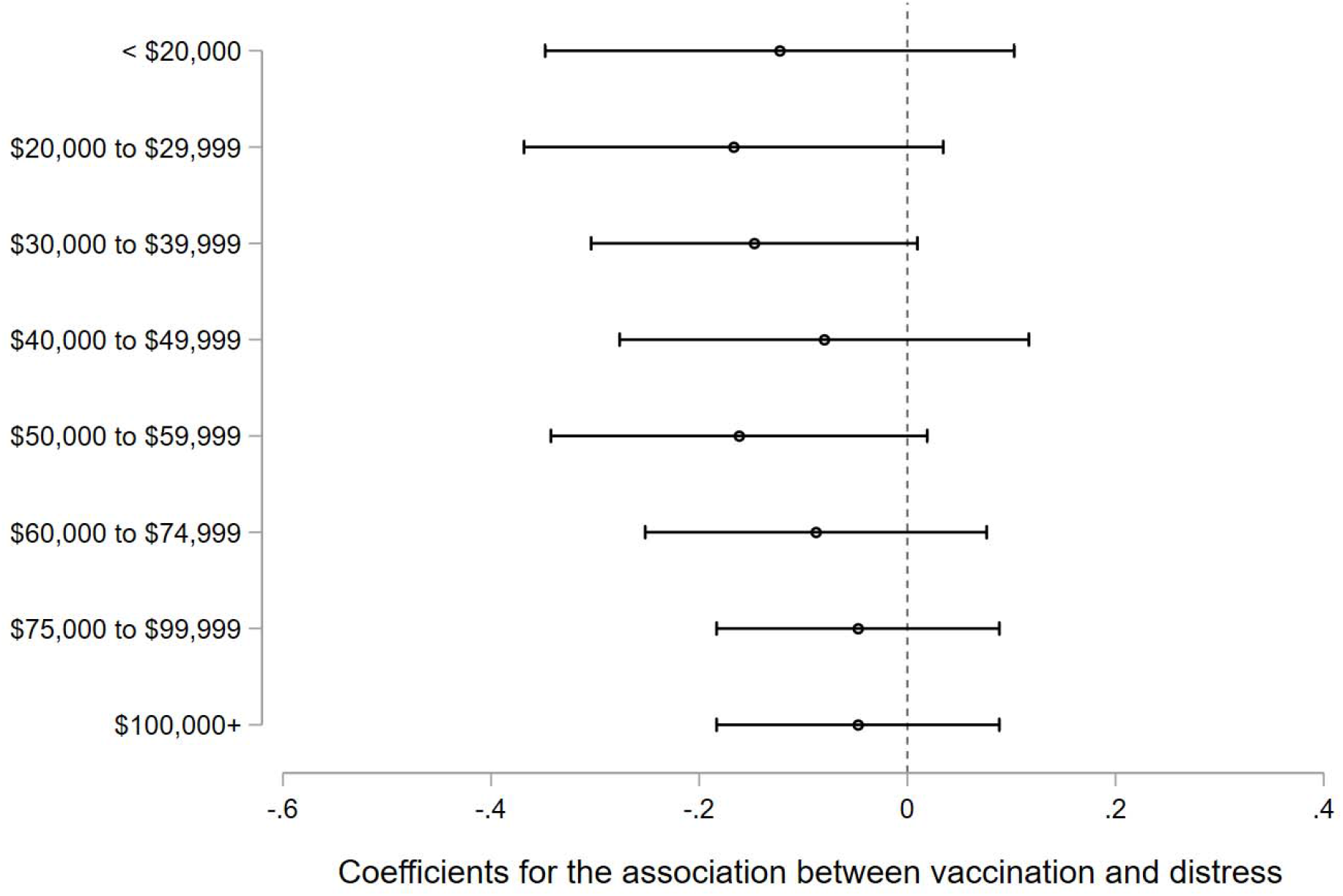
Two-way fixed effects models with psychological distress (PHQ-4) regressed on vaccination status, stratified by household income, April 2020 to June 2021 Notes: Point estimates and 95% confidence intervals are from separate two-way fixed effects models stratified by subgroup. Models control for individual and wave fixed effects, receiving Supplemental Nutrition Assistance Program (SNAP) benefits in the month prior to the survey, whether the respondent received unemployment insurance in the past 14 days, and employment status at the time of the survey. Standard errors are clustered at the individual level.

**Table A1.**
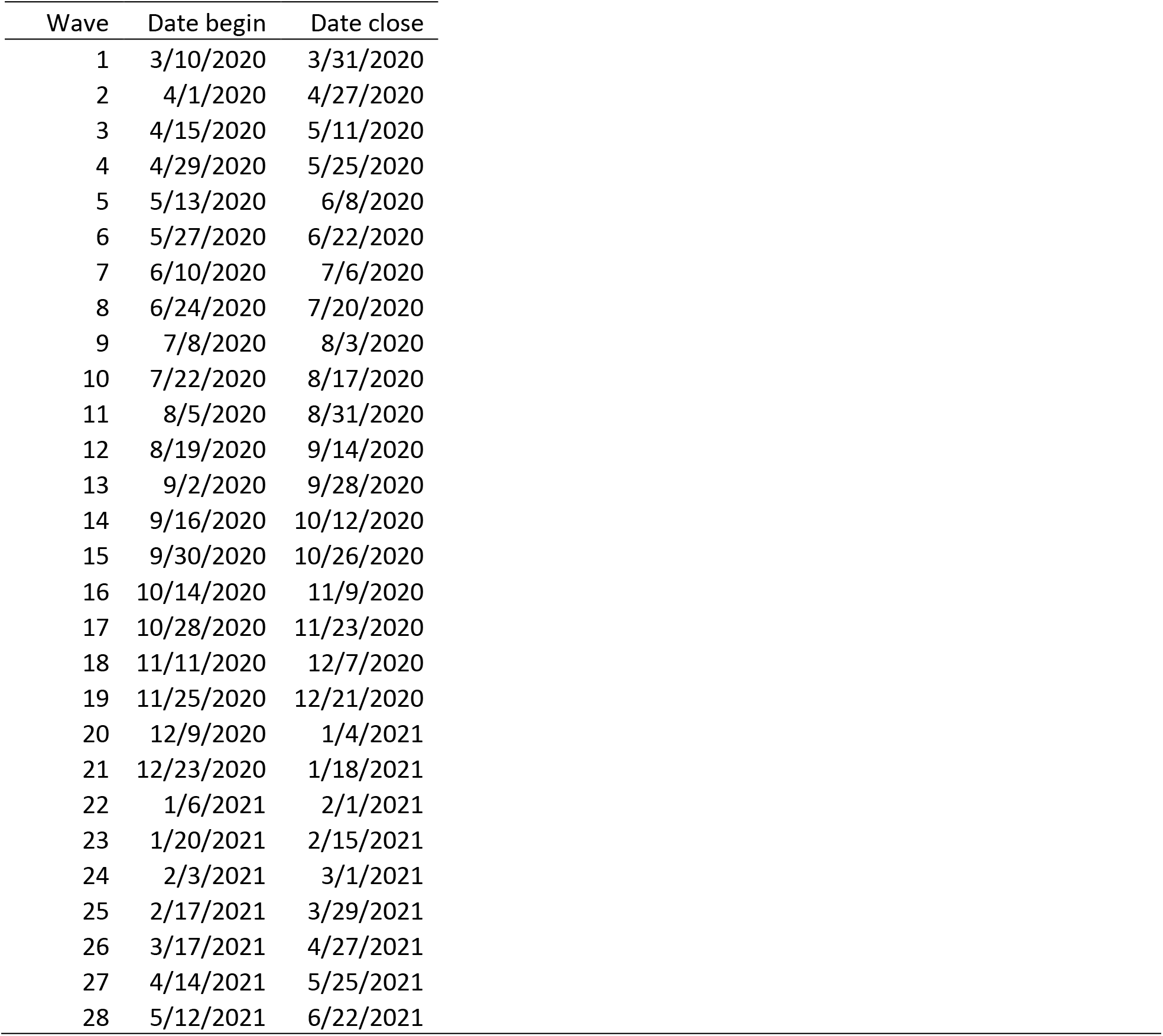
Timing of data collection for each wave of the Understanding Coronavirus in America Notes: Waves 1 and 9 excluded from our TWFE and event study models because core questions were not asked in these periods. New UCA surveys are fielded every two weeks. Each day one fourteenth of participants are invited to take the survey, and participants have two weeks to take the survey – meaning that the total field period for each wave of the survey is 4 weeks and there is overlap between waves. Participants are incentivized to respond to the survey on the day they are invited to participate.

| Wave | Date begin | Date close |
| --- | --- | --- |
| 1 | 3/10/2020 | 3/31/2020 |
| 2 | 4/1/2020 | 4/27/2020 |
| 3 | 4/15/2020 | 5/11/2020 |
| 4 | 4/29/2020 | 5/25/2020 |
| 5 | 5/13/2020 | 6/8/2020 |
| 6 | 5/27/2020 | 6/22/2020 |
| 7 | 6/10/2020 | 7/6/2020 |
| 8 | 6/24/2020 | 7/20/2020 |
| 9 | 7/8/2020 | 8/3/2020 |
| 10 | 7/22/2020 | 8/17/2020 |
| 11 | 8/5/2020 | 8/31/2020 |
| 12 | 8/19/2020 | 9/14/2020 |
| 13 | 9/2/2020 | 9/28/2020 |
| 14 | 9/16/2020 | 10/12/2020 |
| 15 | 9/30/2020 | 10/26/2020 |
| 16 | 10/14/2020 | 11/9/2020 |
| 17 | 10/28/2020 | 11/23/2020 |
| 18 | 11/11/2020 | 12/7/2020 |
| 19 | 11/25/2020 | 12/21/2020 |
| 20 | 12/9/2020 | 1/4/2021 |
| 21 | 12/23/2020 | 1/18/2021 |
| 22 | 1/6/2021 | 2/1/2021 |
| 23 | 1/20/2021 | 2/15/2021 |
| 24 | 2/3/2021 | 3/1/2021 |
| 25 | 2/17/2021 | 3/29/2021 |
| 26 | 3/17/2021 | 4/27/2021 |
| 27 | 4/14/2021 | 5/25/2021 |
| 28 | 5/12/2021 | 6/22/2021 |

**Table 2A:**
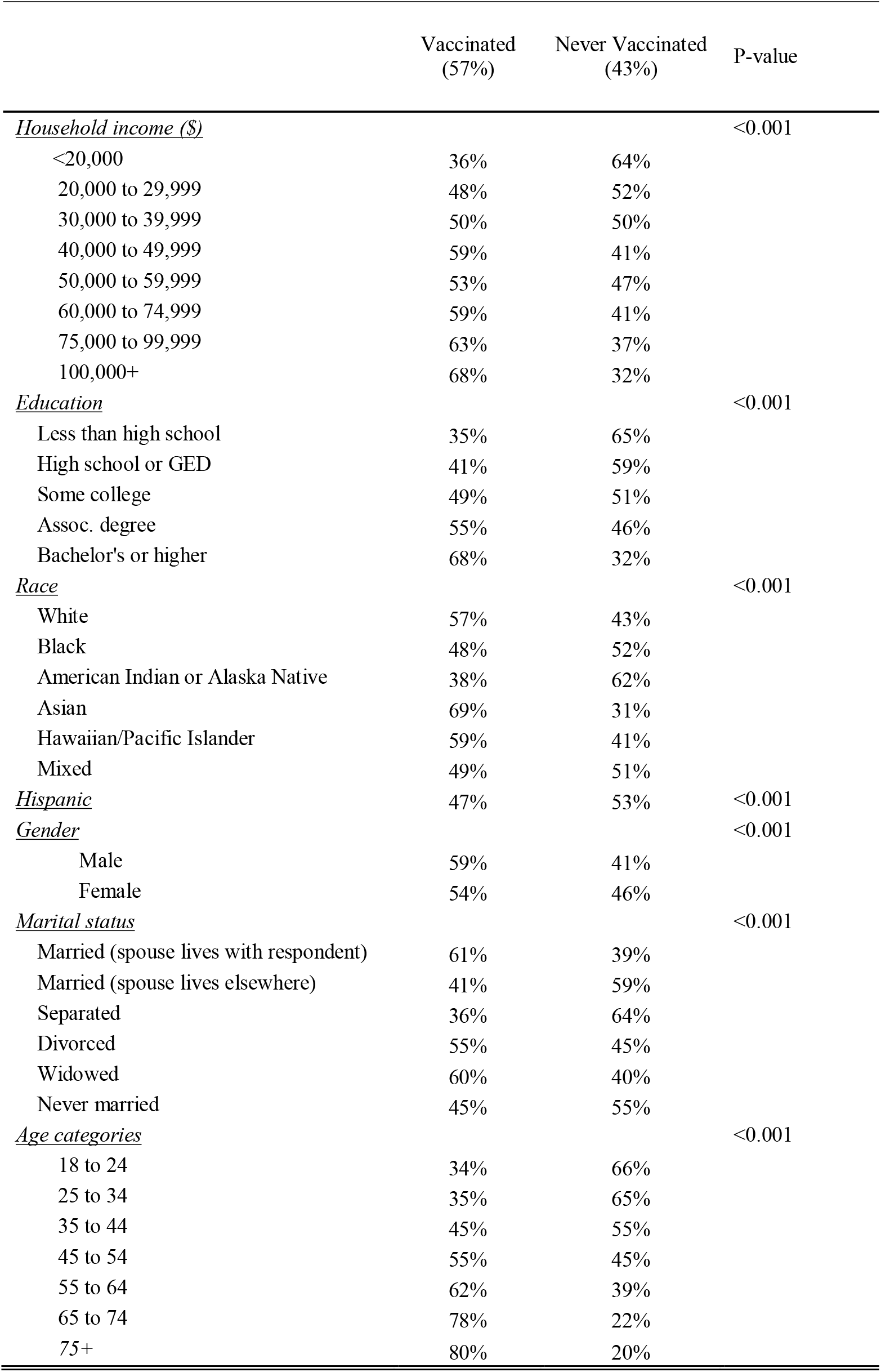
Characteristics of sample by vaccination status Note: Characteristics are unweighted and based on the first observation for each participant in the sample. P-values are from chi-square tests of the bivariate association between the demographic group and a dichotomous variable that equals 1 if the respondent indicates being vaccinated in any wave of the survey and 0 if never vaccinated.

|  | Vaccinated<br>(57%) | Never Vaccinated<br>(43%) | P-value |
| --- | --- | --- | --- |
| <u>Household income (\$)</u> | | | <0.001 |
| <20,000 | 36% | 64% |  |
| 20,000 to 29,999 | 48% | 52% |  |
| 30,000 to 39,999 | 50% | 50% |  |
| 40,000 to 49,999 | 59% | 41% |  |
| 50,000 to 59,999 | 53% | 47% |  |
| 60,000 to 74,999 | 59% | 41% |  |
| 75,000 to 99,999 | 63% | 37% |  |
| 100,000+ | 68% | 32% |  |
| <u>Education</u> |  |  | <0.001 |
| Less than high school | 35% | 65% |  |
| High school or GED | 41% | 59% |  |
| Some college | 49% | 51% |  |
| Assoc. degree | 55% | 46% |  |
| Bachelor's or higher | 68% | 32% |  |
| <u>Race</u> |  |  | <0.001 |
| White | 57% | 43% |  |
| Black | 48% | 52% |  |
| American Indian or Alaska Native | 38% | 62% |  |
| Asian | 69% | 31% |  |
| Hawaiian/Pacific Islander | 59% | 41% |  |
| Mixed | 49% | 51% |  |
| <u>Hispanic</u> | 47% | 53% | <0.001 |
| <u>Gender</u> |  |  | <0.001 |
| Male | 59% | 41% |  |
| Female | 54% | 46% |  |
| <u>Marital status</u> |  |  | <0.001 |
| Married (spouse lives with respondent) | 61% | 39% |  |
| Married (spouse lives elsewhere) | 41% | 59% |  |
| Separated | 36% | 64% |  |
| Divorced | 55% | 45% |  |
| Widowed | 60% | 40% |  |
| Never married | 45% | 55% |  |
| <u>Age categories</u> |  |  | <0.001 |
| 18 to 24 | 34% | 66% |  |
| 25 to 34 | 35% | 65% |  |
| 35 to 44 | 45% | 55% |  |
| 45 to 54 | 55% | 45% |  |
| 55 to 64 | 62% | 39% |  |
| 65 to 74 | 78% | 22% |  |
| 75+ | 80% | 20% |  |

**Table 3A.**
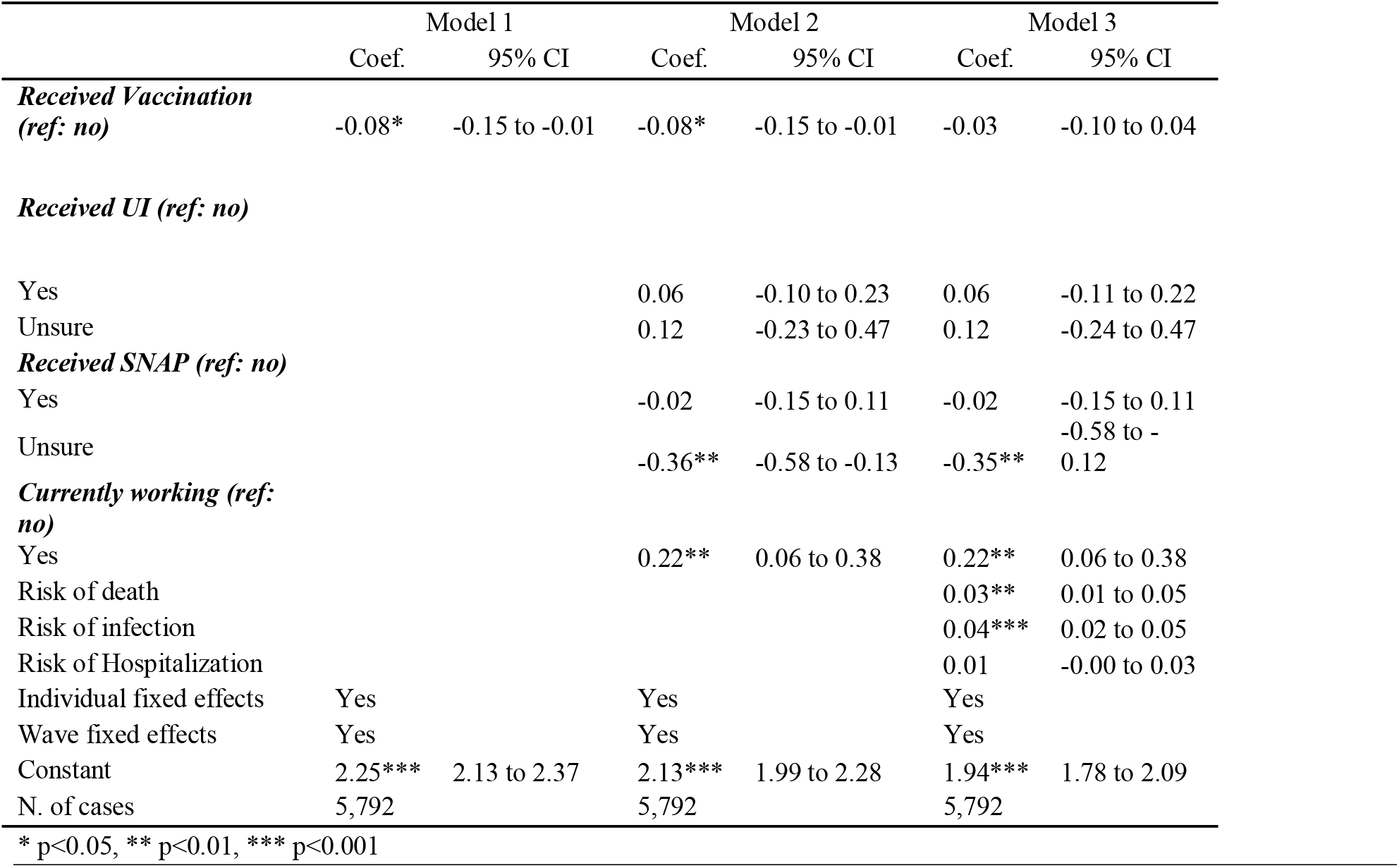
Two-way fixed effects models with psychological distress (PHQ-4) regressed on vaccination status and perceived risk factors using sample weights, April 2020 to June 2021 (N= 5,792)

**Table 4A.** Two-way fixed effects models with psychological distress (PHQ-4) regressed on vaccination status and perceived risk factors among respondents aged 65 and above, April 2020 to June 2021 (N= 1,473)

|  | Model 1 |  | Model 2 |  | Model 3 |  |
| --- | --- | --- | --- | --- | --- | --- |
|  | Coef. | 95% CI | Coef. | 95% CI | Coef. | 95% CI |
| ***Received Vaccination (ref: no)*** | -0.10* | -0.18 to -0.01 | -0.09* | -0.18 to -0.01 | -0.05 | -0.14 to 0.03 |
| ***Received UI (ref: no)*** |  |  |  |  |  |  |
| Yes |  |  | 0.14 | -0.04 to 0.33 | 0.14 | -0.04 to 0.33 |
| Unsure |  |  | 0.43 | -0.30 to 1.16 | 0.42 | -0.30 to 1.14 |
| ***Received SNAP (ref: no)*** |  |  |  |  |  |  |
| Yes |  |  | -0.03 | -0.29 to 0.23 | -0.03 | -0.28 to 0.23 |
| Unsure |  |  | 0.00 | -0.34 to 0.33 | 0.01 | -0.33 to 0.35 |
| ***Currently working (ref: no)*** |  |  |  |  |  |  |
| Yes |  |  |  |  |  |  |
| Risk of death |  |  |  |  | 0.01 | -0.01 to 0.03 |
| Risk of infection |  |  |  |  | 0.03*** | 0.01 to 0.04 |
| Risk of Hospitalization |  |  |  |  | 0.01* | 0.00 to 0.03 |
| Individual fixed effects | Yes |  | Yes |  | Yes |  |
| Wave fixed effects | Yes |  | Yes |  | Yes |  |
| Constant | 1.47*** | 1.37 to 1.56 | 1.45*** | 1.35 to 1.55 | 1.31*** | 1.19 to 1.43 |
| N. of cases | 1,473 |  | 1,473 |  | 1,473 |  |

\* p<0.05, \*\* p<0.01, \*\*\* p<0.001

**Table 5A.** Two-way fixed effects models with severe psychological distress (PHQ-4 scores ≥ 9) regressed on vaccination status and perceived risk factors, April 2020 to June 2021 (N= 5,792) Notes: Coefficients are from linear probability models. Severe distress is coded 1 if PHQ-4 scores are equal to or greater than 9 and coded 0 for scores below 9.

|  | Model 1 |  | Model 2 |  | Model 3 |  |
| --- | --- | --- | --- | --- | --- | --- |
|  | Coef. | 95% CI | Coef. | 95% CI | Coef. | 95% CI |
| ***Received Vaccination (ref: no)*** | -0.006* | -0.011,-0.001 | -0.006* | -0.011,-0.001 | -0.003 | -0.008,0.002 |
| ***Received UI (ref: no)*** |  |  |  |  |  |  |
| Yes |  |  | -0.004 | -0.014,0.005 | -0.004 | -0.014 to 0.005 |
| Unsure |  |  | -0.008 | -0.033,0.016 | -0.009 | -0.033 to 0.016 |
| ***Received SNAP (ref: no)*** |  |  |  |  |  |  |
| Yes |  |  | -0.005 | -0.017 to 0.007 | -0.005 | -0.017 to 0.007 |
| Unsure |  |  | -0.014 | -0.031 to 0.002 | -0.014 | -0.030 to 0.002 |
| ***Currently working (ref: no)*** |  |  |  |  |  |  |
| Yes |  |  | 0.008 | -0.004 to 0.019 | 0.007 | -0.004 to 0.019 |
| Risk of death |  |  |  |  | 0.001* | 0.000 to 0.003 |
| Risk of infection |  |  |  |  | 0.002*** | 0.001 to 0.003 |
| Risk of Hospitalization |  |  |  |  | 0.000 | -0.001 to 0.001 |
| Individual fixed effects |  |  |  |  |  |  |
| Wave fixed effects |  |  |  |  |  |  |
| Constant | 0.064*** | 0.055 to 0.072 | 0.061*** | 0.050 to 0.071 | 0.051*** | 0.040 to 0.063 |
| N. of cases | 5,792 |  | 5,792 |  | 5,792 |  |

\* p<0.05, \*\* p<0.01, \*\*\* p<0.001

**Table 6A.**
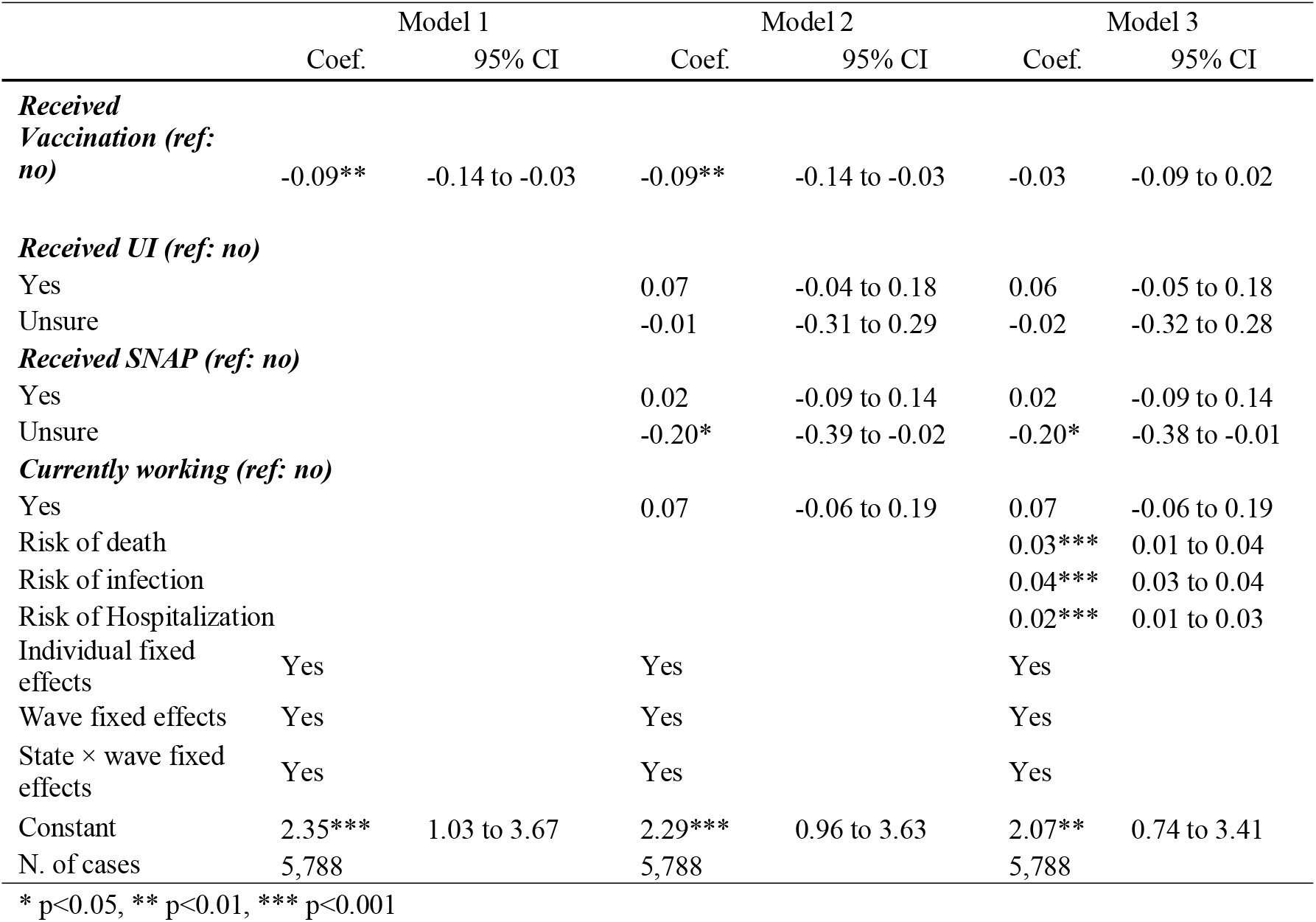
Two-way fixed effects models with severe psychological distress (PHQ-4 scores 9 and above) regressed on vaccination status and perceived risk factors adjusting for state-by-wave fixed effects, April 2020 to June 2021 (N= 5,788)

